# Atovaquone/Proguanil Use and Zoster Vaccination Are Associated with Reduced Alzheimer’s Disease Risk in Two Cohorts: Implications for a Latent *Toxoplasma gondii* Mechanism

**DOI:** 10.1101/2025.03.07.25323566

**Authors:** Ariel Israel, Abraham Weizman, Sarah Israel, Shai Ashkenazi, Shlomo Vinker, Eli Magen, Eugene Merzon

## Abstract

**INTRODUCTION:** Identifying modifiable risk factors for Alzheimer’s disease (AD) may shed light on novel mechanisms and inform prevention strategies. Increasing evidence suggests that latent pathogens may contribute to AD pathogenesis via chronic neuroinflammation.

**METHODS:** We conducted a large-scale, dual-cohort study to identify exposures associated with reduced Alzheimer’s disease (AD) risk. In a national Israeli cohort (Leumit Health Services; 2004-2024), we analyzed 9,124 AD patients and 18,248 matched controls. We systematically screened medication exposures in the matched cohort for associations with significantly reduced AD risk (OR < 0.5, FDR < 0.05). To account for potential residual confounding, we applied conditional logistic regression models adjusted for age, sex, socioeconomic status, and relevant comorbidities.

Findings were independently validated in the U.S.-based TriNetX network, which includes electronic health records from over 120 million patients across 69 healthcare organizations. Propensity score-matched Cox proportional hazards models were used to estimate hazard ratios (HRs) for dementia incidence across stratified age groups.

**RESULTS:** Atovaquone/proguanil (Ato/Pro), an antiprotozoal agent active against *Toxoplasma gondii*, was strongly associated with reduced AD risk in both cohorts (LHS: OR 0.36 [95% CI, 0.20-0.61]; TriNetX: HRs 0.34-0.51, *p* = 10^-17^ to 10^-40^ across age groups 50-59, 60-69, and 70-79). Both recombinant and live attenuated varicella-zoster virus (VZV) vaccines were also significantly protective (ORs 0.16-0.37), and *T. gondii* seropositivity was associated with a 2.43-fold increased risk of dementia (*p* = 0.0013). Notably, Ato/Pro’s protective effect was more pronounced in individuals without prior VZV vaccination (HR 0.51 [0.43-0.59]) compared to vaccinated individuals (HR 0.71 [0.59-0.85]).

**DISCUSSION:** This dual-cohort study - spanning over 120 million patients across two nations - demonstrates strong and reproducible associations linking Ato/Pro use and VZV vaccination to reduced AD risk. The findings support a mechanistic model in which latent *T. gondii* infection, potentially reactivated by herpesvirus co-infection may contribute to AD pathogenesis. Ato/Pro may protect by eliminating or suppressing *T. gondii*, while VZV vaccination may reduce viral triggers of parasite reactivation. These results point to novel preventive strategies and reinforce the infectious hypothesis of Alzheimer’s disease.

## Background

Alzheimer’s disease (AD) is the most common cause of dementia, characterized by progressive cognitive decline and memory impairment. Globally, over 50 million individuals are affected by dementia, with AD accounting for 60-70% of these cases. This prevalence is projected to triple by 2050, reaching an estimated 152 million cases.(Contador et al., 2024)

The etiology of AD is multifactorial, involving both genetic and environmental factors. Approximately 70% of the risk can be attributed to genetics, notably the presence of the apolipoprotein E ε4 allele.(Silva et al., 2019) However, modifiable risk factors also play a significant role. Epidemiological studies have identified associations between AD and various factors, including hypertension, lifestyle, and cerebrovascular diseases. Conversely, protective factors such as higher educational attainment, cognitive engagement, and regular physical activity have been linked to a reduced risk of AD. (Qiu et al., 2009) AD is pathologically characterized by the accumulation of amyloid-beta plaques and neurofibrillary tangles, leading to synaptic dysfunction and neuronal loss. Emerging evidence suggests that pathogen-induced neuroinflammation may exacerbate these pathological features, as infections can trigger chronic inflammatory responses in the brain, potentially accelerating AD progression.(Hensley, 2010)

Recent research has explored the potential protective effects of vaccinations against AD. Immunizations, notably VZV vaccines, have been associated with a decreased risk of developing AD(Pomirchy et al., 2025; Taquet et al., 2024). The proposed mechanisms include the enhancement of the immune system’s ability to combat infections that may contribute to neuroinflammation and subsequent neurodegeneration.(Ukraintseva et al., 2024) Vaccination may also modulate immune responses that affect amyloid-beta clearance, reducing AD pathology.

The role of infections in AD pathogenesis and other neurodegenerative processes has garnered increasing attention. Several microbial taxa, including *Porphyromonas gingivalis(Dominy et al., 2019)*, and herpesviruses(Piotrowski et al., 2023; Readhead et al., 2018), have been implicated in AD pathology through inflammatory and neurotoxic mechanisms. Moreover, antibodies against *Toxoplasma gondii*, a common parasitic infection, have been found at higher rates in individuals with neuropsychiatric disorders such as schizophrenia(Kusbeci et al., 2011; Sutterland et al., 2015; Torrey et al., 2012), and emerging evidence suggests a possible link to AD(Kusbeci et al., 2011; Wang et al., 2024; Yang et al., 2021). Chronic infections may contribute to sustained neuroinflammation, leading to neuronal damage, synaptic dysfunction, and cognitive decline. Understanding the interplay between the microbiome, chronic infections, and neurodegeneration is critical for identifying novel preventive and therapeutic strategies for AD.(Lathe et al., 2023)

In this study, we leveraged electronic health records (EHRs) from a national health organization in Israel to systematically investigate whether individuals who developed AD over the past two decades exhibited distinct patterns of previous medication use. Identifying significant reductions in AD occurrence among patients who had received prior antimicrobial treatments may provide insights into the role of targeted microorganisms in the pathogenic processes leading to AD development.

## Methods

### Study design

The first stage of this research was conducted as a retrospective cohort study in Leumit Health Services (LHS), one of the four nationwide health providers in Israel, providing comprehensive healthcare services to approximately 730,000 members. All Israeli citizens are entitled for comprehensive health insurance and receive a standardized package of health services and medications, as defined by the national “Health Basket” committee. LHS operates a centralized EHR system, with over two decades of meticulously maintained information on patient demographics, medical diagnoses, healthcare encounters, laboratory test results, and records of prescribed and purchased medications. Diagnoses are documented during medical encounters by the treating physicians using the International Classification of Diseases (ICD). Diagnoses can be marked as chronic when they pertain to a chronic condition, and these can be updated or closed by the treating physicians during subsequent patient encounters. The reliability of these chronic diagnosis records in our registry has been previously validated, demonstrating high accuracy.(Israel et al., 2021)

Eligibility included all LHS members with active membership between 2003 and 2024. Data were extracted from the LHS central data warehouse, covering diagnoses, laboratory test results, and medication purchases up to December 31, 2024. Structured query language (SQL) and Python scripts automated data retrieval. Socioeconomic status was determined using geocoded residential addresses, classified on a scale of 1 (lowest) to 20 (highest) based on the Points Location Intelligence® database, which strongly correlates with official socioeconomic indicators. Geodemographic classification of the general population, Ultra-Orthodox Jewish, and Arab communities was performed using validated methodologies from prior studies.

### LHS cohort definition

The study included AD patients diagnosed between ages 40 and 89, matched 2:1 with controls who had no dementia diagnosis. To minimize misclassification, individuals with cerebrovascular accident, schizophrenia, Down syndrome, or Parkinson’s disease were excluded. AD cases were identified based on ICD-10 “F00” or ICD-9 “331” codes, with the earliest recorded diagnosis as the index date. Controls were matched to cases by gender, ethnic group (general population, Jewish Ultra-Orthodox, or Arab), socioeconomic status, and year of LHS enrollment. Matching prioritized individuals with the closest birth date to the AD patient, ensuring no duplication.

### TriNetX Validation Cohort

Validation analyses were conducted using TriNetX, a federated research network with EHR data from 69 US healthcare organizations and over 120 million patients. The validation cohorts included individuals aged 65 with no prior dementia diagnosis. Cases were defined as those who had purchased the medication of interest, with controls selected from those who had not. Propensity score matching (PSM) was applied to balance age, ethnicity, race, smoking status, diabetes diagnosis, BMI, and HbA1c levels at medication initiation. Dementia incidence was assessed using Cox proportional hazards models, based on the combined occurrence of ICD-10 codes G30 (Alzheimer’s disease) or F03 (unspecified dementia). Follow-up began 30 days post-index date with no predefined end limit. P-values were calculated using the log-rank test. Using the same methodology, we compared dementia incidence in individuals with past *T. gondii*-positive IgG or IgM serology vs. those with negative serology.

### Data processing

Data from LHS were extracted from EHRs and processed using Python 3.11 scripts and T-SQL queries developed by the Leumit Research Institute. Patient identifiers were removed and replaced with study-specific codes to ensure anonymity. Medication use was assessed based on purchase records up to 10 years before the index date, classified by Anatomical Therapeutic Chemical (ATC) codes.

### Statistical analyses

Analyses of LHS data were conducted using R version 4.4. Fisher’s exact test was used for categorical variables and two-tailed t-tests for continuous variables. Conditional logistic regression models assessed associations between AD and medication use, adjusting for age, gender, smoking status, socioeconomic status, physician visits, pregnancy history, and health worker status.

### Ethics Statement

Ethics approval was granted by the Leumit Health Services Institutional Review Board (LEU-0001-24), with a waiver of informed consent due to retrospective, anonymized data analysis.

## Results

### The study cohort

Table 1 summarizes the demographic and clinical characteristics of 9,124 AD patients and 18,248 controls in the LHS cohort. Due to the matching process, age and gender distributions were very similar (60.7% female, mean age 76.8 ± 8.0 years). AD patients had lower weight (71.5 ± 15.0 vs. 74.3 ± 15.1 kg), height (161 ± 12 vs. 162 ± 10 cm), and BMI. They were more likely to be smokers (OR 1.14 [95% CI, 1.09-1.20]) and had a higher prevalence of diabetes (OR 1.32 [95% CI, 1.25-1.39]), with elevated fasting glucose and hemoglobin A1c levels.

**Table 1.** Demographic and clinical characteristics of the study cohort at index date.

|  |  | **AD**<br>**N=9,124** | **Control**<br>**N=18,248** | **P-**<br>**value** | **Odds Ratio** | **SMD** |
| --- | --- | --- | --- | --- | --- | --- |
| Gender | Female | 5,536 (60.7 %) | 11,072 (60.7 %) | 1.000 | 1 |  |
|  | Male | 3,588 (39.3 %) | 7,176 (39.3 %) | 1.000 | 1 |  |
| Age (years) |  | 76.8 ± 8.0 | 76.7 ± 8.0 | 0.268 |  | 0.014 |
| Age category | 40-49 | 93 (1.02 %) | 184 (1.01 %) | 0.949 | 1.01 [0.78-1.31] |  |
|  | 50-59 | 237 (2.60 %) | 487 (2.67 %) | 0.749 | 0.97 [0.83-1.14] |  |
|  | 60-69 | 1,070 (11.73 %) | 2,180 (11.95 %) | 0.606 | 0.98 [0.90-1.06] |  |
|  | 70-79 | 3,827 (41.94 %) | 7,729 (42.36 %) | 0.525 | 0.98 [0.93-1.03] |  |
|  | 80-89 | 3,897 (42.71 %) | 7,668 (42.02 %) | 0.048 | 1.02 [1.01-1.15] |  |
| Weight (kg) |  | 71.5 ± 15.0 | 74.3 ± 15.1 | <0.001 |  | -0.189 |
|  | missing | 73 (2.23 %) | 2,280 (6.97 %) |  |  |  |
| Height (cm) |  | 161 ± 12 | 162 ± 10 | <0.001 |  | -0.093 |
|  | missing | 122 (3.73 %) | 3,248 (9.92 %) |  |  |  |
| Body Mass Index (kg/m2) |  | 27.8 ± 5.5 | 28.5 ± 5.2 | <0.001 |  | -0.131 |
|  | missing | 1,775 (19.5 %) | 2,672 (14.6 %) |  |  |  |
| BP systolic (mmHg) |  | 134 ± 24 | 135 ± 19 | <0.001 |  | -0.051 |
|  | missing | 100 (1.10 %) | 282 (1.55 %) |  |  |  |
| BP diastolic (mmHg) |  | 75.6 ± 10.1 | 76.0 ± 9.6 | <0.001 |  | -0.043 |
|  | missing | 104 (1.14 %) | 291 (1.59 %) |  |  |  |
| Smoking status | Non-smoker | 2,127 (23.6 %) | 4,564 (25.4 %) | 0.001 | 0.91 [0.85-0.96] |  |
|  | Past smoker | 2,573 (28.5 %) | 5,396 (30.0 %) | 0.010 | 0.93 [0.88-0.98] |  |
|  | Smoker | 4,320 (47.9 %) | 7,997 (44.5 %) | <0.001 | 1.14 [1.09-1.20] |  |
|  | missing | 104 (1.1 %) | 291 (1.6 %) | 0.003 | 0.71 [0.56-0.89] |  |
| Diabetes Mellitus |  | 3,335 (36.6 %) | 5,545 (30.4 %) | <0.001 | 1.32 [1.25-1.39] |  |
| eGFR MDRD (mL/min/1.73m2) |  | 72.7 ± 26.8 | 72.1 ± 22.8 | 0.080 |  | 0.023 |
|  | missing | 152 (1.67 %) | 539 (2.95 %) |  |  |  |
| Glucose (mg/dL) |  | 114 ± 42 | 110 ± 31 | <0.001 |  | 0.122 |
|  | missing | 157 (1.72 %) | 544 (2.98 %) |  |  |  |
| Hemoglobin A1c (%) |  | 6.32 ± 1.32 | 6.19 ± 1.02 | <0.001 |  | 0.113 |
|  | missing | 1,328 (14.6 %) | 3,382 (18.5 %) |  |  |  |
| Free T3 (pmol/L) |  | 4.03 ± 1.01 | 4.16 ± 0.96 | <0.001 |  | -0.137 |
|  | < 3.5 pmol/L | 760 (24.2 %) | 1,134 (18.4 %) | <0.001 | 1.42 [1.28-1.58] |  |
|  | missing | 6,006 (65.7 %) | 12,112 (66.2 %) |  |  |  |
| Vitamin D (ng/mL) |  | 21.4 ± 11.0 | 22.4 ± 10.9 | <0.001 |  | -0.092 |
|  | < 10 ng/mL | 1,067 (14.7 %) | 1,761 (11.5 %) | <0.001 | 1.32 [1.22-1.44] |  |
|  | missing | 1,891 (20.7 %) | 3,027 (16.6 %) |  |  |  |
| Folic Acid (ng/mL) |  | 8.37 ± 8.37 | 8.28 ± 4.82 | 0.015 |  | 0.015 |
|  | < 2.8 ng/mL | 590 (6.76 %) | 783 (4.67 %) | <0.001 | 1.48 [1.32-1.65] |  |
|  | > 17 ng/mL | 724 (8.30 %) | 1,094 (6.53 %) | <0.001 | 1.30 [1.17-1.43] |  |
|  | missing | 418 (4.57 %) | 1,531 (8.37 %) |  |  |  |
| ALT(GPT) (U/L) |  | 19.2 ± 27.1 | 19.3 ± 17.9 | 0.903 |  | -0.001 |
|  | < 8 U/L | 534 (5.85 %) | 592 (3.28 %) | <0.001 | 1.83 [1.62-2.07] |  |
|  | missing | 19 (0.21 %) | 231 (1.26 %) |  |  |  |

SMD: Standardized Mean Difference (Cohen's D)

Notable laboratory differences included a higher proportion of AD patients with free T3 levels <3.5 pmol/L (OR 1.42 [95% CI, 1.28-1.58]) or severe vitamin D deficiency of <10 ng/mL (OR 1.32 [95% CI, 1.22-1.44]). Folic acid levels showed a bimodal distribution, with AD patients being more likely to have both deficiency (<2.8 ng/mL, OR 1.48 [95% CI, 1.32-1.65]) and excess (>17 ng/mL, OR 1.30 [95% CI, 1.17-1.43]). Although mean ALT levels were comparable, AD patients had a higher prevalence of very low ALT (<8 U/L, OR 1.83 [95% CI, 1.62-2.07]).

### Medication use associated with the risk of AD occurrence

Medication use over the past decade was systematically assessed for associations with AD risk using Fisher’s exact test. Table 2 presents the three medications that met the criteria of OR <0.5 and FDR <5% (Benjamini-Hochberg correction). The two VZV vaccines were significantly associated with lower AD risk: the recombinant VZV vaccine (OR 0.16 [95% CI, 0.05-0.40]) and the live attenuated VZV vaccine (OR 0.37 [95% CI, 0.26-0.52]). Additionally, we observed a significant reduction in AD risk with Atovaquone/Proguanil (Ato/Pro) use, an antiprotozoal used for malaria prophylaxis (OR 0.36 [95% CI, 0.20-0.61]). This combination drug inhibits protozoal mitochondrial function and disrupts folate metabolism.

**Table 2.** Comparison of medication purchases in the 10 years preceding the index date in the AD and control groups of the LHS cohort.

| **Medication** | **AD**<br>**N=9,124** | **Control**<br>**N=18,248** | **Odds Ratio**<br>**[95% CI]** | **adjusted Odds**<br>**Ratio**<br>**[95% CI]** | **adjusted**<br>**p-value** |
| --- | --- | --- | --- | --- | --- |
| Atovaquone + Proguanil (PO) | 17 (0.19%) | 94 (0.51%) | 0.36 [0.20-0.61] | 0.41 [0.24-0.69] | 0.0008 |
| Zoster, Purified antigen vaccine (IM) | 5 (0.05%) | 62 (0.34%) | 0.16 [0.05-0.40] | 0.20 [0.08-0.50] | 0.0006 |
| Zoster, Live attenuated vaccine (SC) | 41 (0.45%) | 218 (1.19%) | 0.37 [0.26-0.52] | 0.39 [0.28-0.55] | <0.0001 |

PO: Per Os; SC: Subcutaneous; IM: Intramuscular
Odds ratio were computed according to Fisher's exact tests performed on the matched cohort
Adjusted odds ratio and p-value, were computed according to a conditional logistic regression model which included all the listed medications and is adjusted for age (years), gender, smoking status, SES category, number of past physician visits, previous pregnancy, and health worker status

To further assess the independent effect of each medication on AD risk, we performed an adjusted multivariable regression analysis incorporating these three medications along with potential confounders. The associations remained robust, with adjusted odds ratios and p-values presented in the two rightmost columns of Table 2.

### Validation in the US based trinetX collaborative network

To validate the associations observed in the national cohort, we conducted an independent analysis using TriNetX, a global federated network comprising electronic health records (EHRs) from over 69 U.S. healthcare organizations and more than 120 million patients.

Results are summarized in Table 3, with selected comparisons illustrated as cumulative incidence plots in Figure 1 (subplot letters correspond to the first column of Table 3). The analysis focused on three age strata - 50-59, 60-69, and 70-79 years - each including individuals who had undergone an ambulatory visit during the study period, which served as the index date.

**Figure 1.**
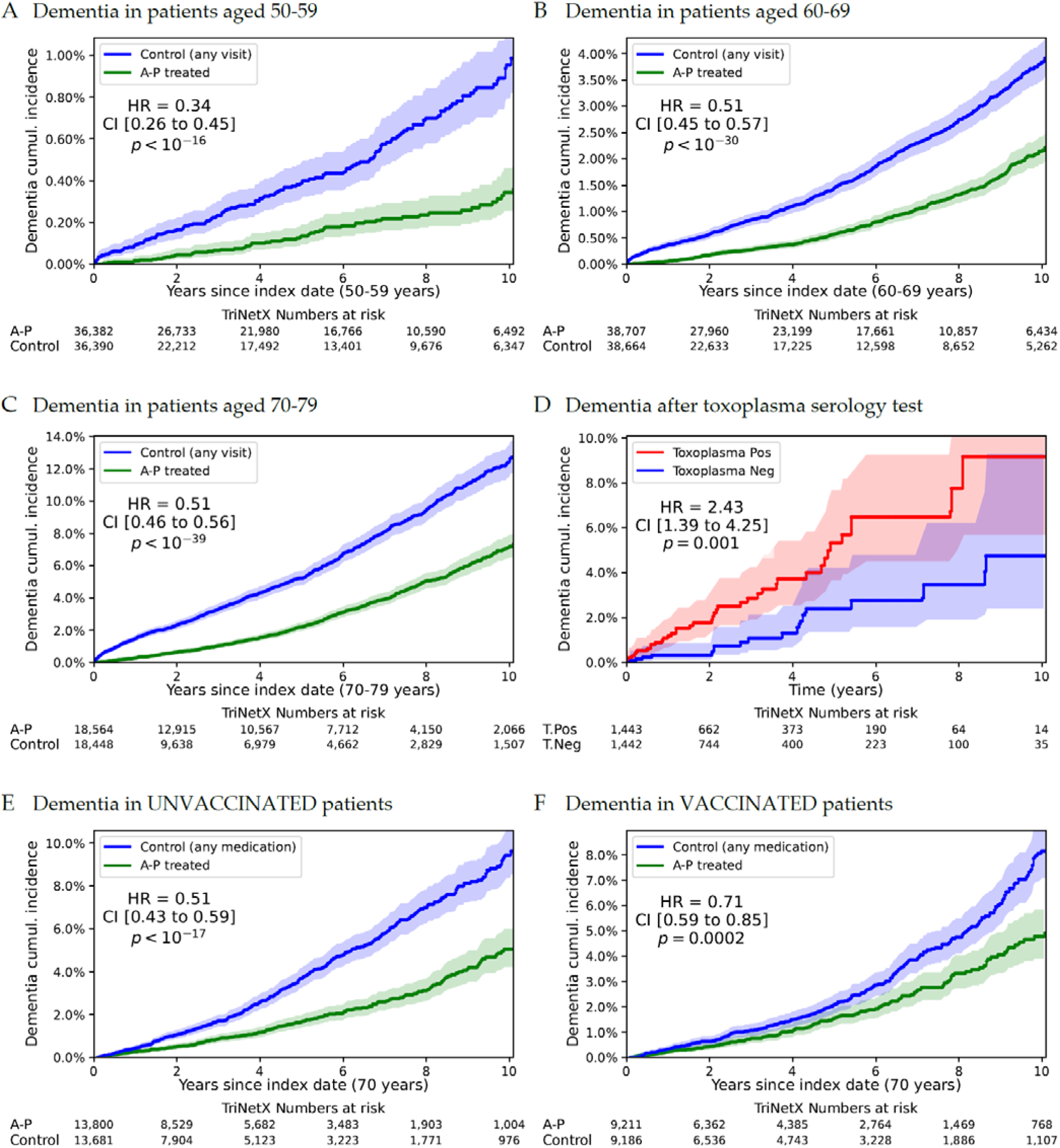
Cumulative incidence of dementia across matched cohorts in TriNetX U.S. data. Panels A-C show reduced dementia incidence among individuals previously treated with atovaquone/proguanil (A+P) compared to matched controls across three age groups. Panel D shows increased dementia incidence among *Toxoplasma gondii* seropositive vs. seronegative individuals. Panels E and F stratify the A+P dementia risk by prior varicella-zoster vaccination status. Shaded bands represent 95% confidence intervals. All cohorts were matched 1:1 on age, gender, race/ethnicity, BMI, diabetes, and smoking status. “Control” refers to individuals without A+P exposure (A-C) or with any matched medication use (E, F).

**Table 3.**
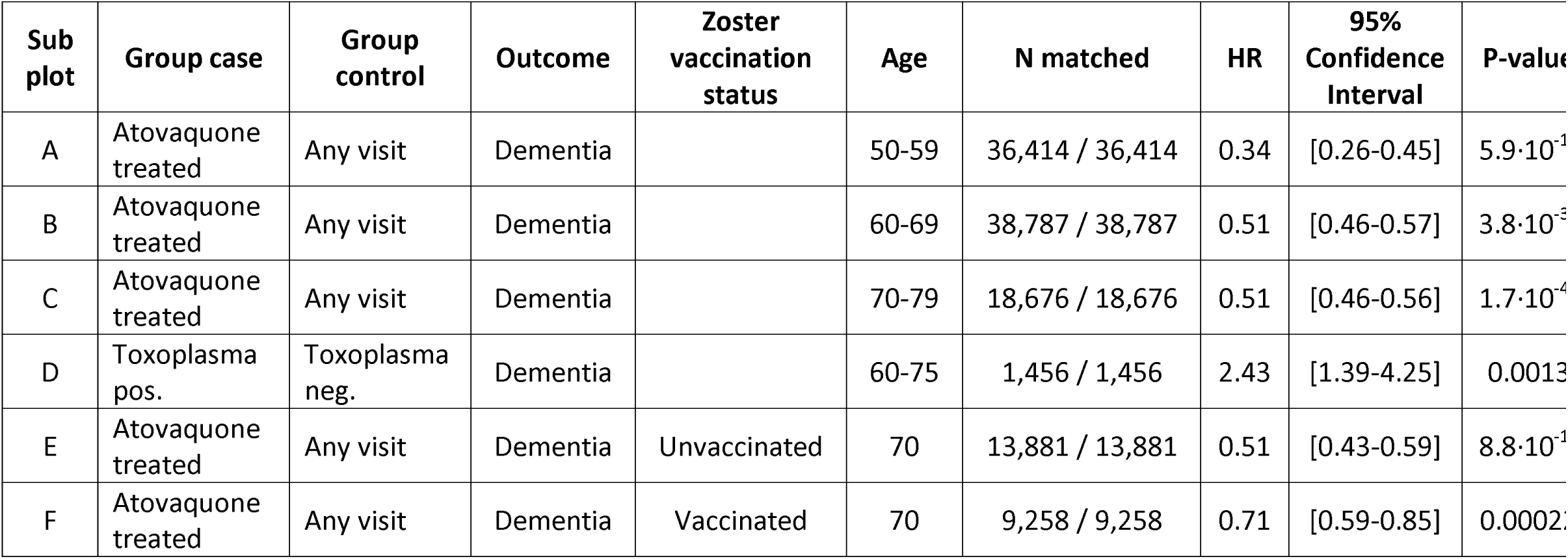
Outcome Comparisons Studies in TriNetX U.S. cohort. Dementia outcome comparisons in matched cohorts from the TriNetX U.S. network. This table summarizes hazard ratios (HR), confidence intervals, and p-values for dementia incidence across multiple matched comparisons. Subplots A-F correspond to specific analyses displayed in Figure 1. Comparisons include patients exposed to atovaquone/proguanil across three age strata (50-59, 60-69, and 70-79), individuals with positive vs. negative *Toxoplasma gondii* serology (ages 60-75), and stratification by zoster vaccination status at age 70. All cohorts were matched 1:1 by propensity scores on demographics and clinical covariates. Dementia was defined by ICD-10 codes G30 or F03.

The Ato/Pro-exposed groups consisted of individuals with a prior documented purchase of atovaquone and proguanil. Control groups included individuals without such exposure. Propensity score matching was applied to balance the cohorts on age, gender, race/ethnicity, diabetes status, body mass index (BMI), and smoking status. Individuals with a previous diagnosis of Alzheimer’s disease or any form of dementia before the index date were excluded from the analysis.

Across all three age strata, Ato/Pro use was consistently associated with a significant reduction in AD risk, closely replicating the findings from our national cohort:

- Ages 50-59: 36,414 matched pairs, HR = 0.34 [95% CI, 0.26-0.45], P=5.9·10^-17^
- ges 60-69: 38,787 matched pairs, HR = 0.51 [95% CI, 0.46-0.57], P=3.8·10^-31^
- Ages 70-79: 18,676 matched pairs, HR = 0.51 [95% CI, 0.46-0.56], P=1.7·10^-40^
- T. gondii IgG/IgM positive vs. negative: 1,456 matched pairs, HR = 2.43 [95% CI, 1.39-4.25], P=0.0013

To assess whether *T. gondii*, a parasite eliminated by Ato/Pro, is linked to dementia, we analyzed individuals under 85 years old with a prior *T. gondii* IgG or IgM test. We compared 1,456 individuals with positive serology with a matched group of individuals with negative serology and found a significantly higher incidence of dementia in the *T. gondii* positive group (HR 2.43, p=0.001; Fig. 1B). These findings independently confirm the inverse association between Ato/Pro use and AD risk, and support the hypothesis that *T. gondii* may contribute to dementia pathogenesis.

We further examined whether VZV vaccination influenced the protective effect of Ato/Pro by stratifying individuals aged ≥70 based on their vaccination status. Among unvaccinated individuals (no prior VZV vaccine purchase), Ato/Pro use was associated with a greater and more significant reduction in AD risk (HR 0.51 [95% CI, 0.43-0.59]; Fig. 1C) compared to vaccinated individuals, who had received a VZV vaccine before the index date (HR 0.71 [95% CI, 0.59-0.85]; Fig. 1D). These findings suggest that Ato/Pro’s protective effect against AD may be more pronounced in individuals who had not received a VZV vaccine.

## Discussion

This study identifies a novel and robust association between prior atovaquone/proguanil (Ato/Pro) use and a significantly reduced risk of Alzheimer’s disease (AD). Conducted within a nationwide health provider over a 20-year period, our large-scale retrospective analysis included 9,124 AD patients and 18,248 matched controls. The inverse association between Ato/Pro use and AD risk was both statistically significant and clinically meaningful, suggesting a previously unrecognized protective factor against AD. Interestingly, our study also identified the protective effect of both the live attenuated and recombinant varicella-zoster virus (VZV) vaccines reported recently by other studies(Pomirchy et al., 2025; Taquet et al., 2024).

To minimize the possibility of reverse causation, we excluded individuals with any prior diagnosis of dementia or prodromal neurodegenerative conditions at baseline. Additionally, we ensured that all medication exposures occurred at least one month prior to the index date to establish proper temporal sequencing.

Notably, we did not observe a protective association with other medications known to have some anti-*Toxoplasma* activity, such as oral clindamycin. However, this may reflect differences in clinical usage patterns. While Ato/Pro is typically prescribed for extended durations - often spanning several weeks, including pre- and post-travel periods - clindamycin is generally avoided in our health system due to concerns about *Clostridioides difficile* infection risk. When used, it is prescribed almost exclusively to penicillin-allergic patients, typically in short courses and at low doses, which are unlikely to achieve effective anti-*Toxoplasma* activity. This distinction in dosing and treatment context may explain the lack of observed protective effect for clindamycin.

To validate these findings, we independently analyzed data from TriNetX, a U.S.-based global network encompassing electronic health records from over 120 million patients. Using established analytical frameworks and propensity score matching, we observed a consistent and reproducible protective association between Ato/Pro use and reduced dementia risk across three distinct age groups (50-59, 60-69, and 70-79 years). The consistency of this association across age strata, combined with extremely low p-values (ranging from 10-¹ to 10-), reinforces the strength and reproducibility of the observed effect.

Importantly, Kaplan-Meier analyses revealed a sustained divergence in cumulative dementia incidence curves extending over more than a decade following Ato/Pro exposure. This long-term protective effect strengthens the case for a causal relationship, with the elimination of a persistent pathogen - most plausibly *Toxoplasma gondii* - representing the most likely underlying mechanism. Consistent with the LHS cohort design, we excluded all individuals with a prior diagnosis of dementia in the TriNetX analyses to minimize confounding by indication and reduce the likelihood of including individuals with early or undiagnosed cognitive decline.

We also found that *Toxoplasma gondii* seropositivity was significantly associated with subsequent dementia development, with most diagnoses occurring many years after the serology was performed. This temporal separation argues strongly against reverse causation - particularly as *T. gondii* serologic testing is not part of routine dementia evaluation - and further supports a potential role for chronic, latent toxoplasmosis in the pathogenesis of Alzheimer’s disease.

Interestingly, the protective effect of Ato/Pro was stronger in individuals who had not received a VZV vaccine compared to those who had, suggesting a possible interaction between herpesvirus infection and *T. gondii* reactivation in driving neurodegenerative processes. These observations point to a convergent pathogenic model in which latent *T. gondii* infection, possibly reactivated by herpesvirus superinfection, contributes to chronic neuroinflammation and AD development. In this framework, both Ato/Pro and VZV vaccination act through distinct but complementary mechanisms to reduce AD risk.

Based on our findings, we propose that latent *T. gondii* may contribute to AD pathogenesis. Specifically, we hypothesize that herpesvirus infection may trigger *T. gondii* reactivation, leading to neuroinflammation that drives AD pathology in genetically or immunologically susceptible individuals.

### Toxoplasma gondii

*T. gondii* is a ubiquitous protozoan parasite commonly found in the environment, with humans frequently acquiring asymptomatic, lifelong infections. While generally latent, *T. gondii* has been implicated in a range of neurological and psychiatric disorders(Flegr, 2007; Henriquez et al., 2009), including schizophrenia(Torrey et al., 2012), depression(Hlaváčová et al., 2021), and epilepsy(Ngoungou et al., 2015). Epidemiological studies have reported associations between *T. gondii* seropositivity and an increased risk of neuropsychiatric conditions, raising the possibility that chronic infection may contribute to neuroinflammation and progressive neuronal damage.

Findings on the association between *T. gondii* and AD have been inconsistent. While some cohorts found no significant link(Wyman et al., 2017), recent cohort studies reported significant associations between *T. gondii* infection and dementia risk.(Wang et al., 2024; Yang et al., 2021) Additionally, an NHANES analysis found *T. gondii* seropositivity correlated with lower cognitive function scores, further suggesting neurological impacts of chronic infection(Wiener et al., 2020).

Since *T. gondii* shares mitochondrial and folate metabolism pathways with other protozoa, Atovaquone is effective against it and is used with pyrimethamine as a DHFR inhibitor in toxoplasmosis treatment.(KOVACS, 1992)

Our systematic screening revealed significant AD protection also from both varicella zoster vaccines, likely by preventing herpesvirus infections in neurons, prompting a hypothesis linking herpesvirus reactivation, *T. gondii*, and AD pathology. We propose that in susceptible individuals, herpesvirus infection triggers reactivation of latent *T. gondii*, releasing bradyzoites that could then spread to adjacent neurons, inducing chronic neuroinflammation contributing to amyloid plaques and neurodegeneration. This is supported by Ato/Pro reducing AD risk much more strongly in patients who had not been vaccinated for VZV (HR=0.51, P<0.0001), suggesting both *T. gondii* and herpesvirus (likely VZV) drive pathogenesis. Herpesvirus signatures detected across multiple AD cohorts reinforce this model.(Readhead et al., 2018)

Neuronal cells, being quiescent and metabolically constrained, severely limit *Toxoplasma gondii*’s ability to replicate. Consequently, primary infection is typically contained within the CNS through an immune response that includes microglial activation and cytokine production. This response forces the parasite into its dormant bradyzoite stage, allowing *T. gondii* to evade immune surveillance and persist within host cells. Herpesvirus superinfection of *T. gondii*-harboring cells may disrupt this equilibrium. Viral replication-induced cytolysis can lead to the lysis of infected cells, releasing *T. gondii* into the extracellular space. The resulting neuroinflammatory response—characterized by cytokine release and increased blood-brain barrier (BBB) permeability—may facilitate further invasion of the parasite into adjacent neuronal and glial cells. This cascade of events could amplify neuroinflammation, neuronal damage, and neurodegeneration, ultimately contributing to AD pathology.

### Liver Dysfunction and Immune Dysregulation

A significant proportion of AD patients from the LHS cohort exhibited very low alanine aminotransferase (ALT) levels, suggestive of hepatic dysfunction. The liver plays a key role in immune surveillance and pathogen clearance, filtering pathogens via Kupffer cells, natural killer (NK) cells, and T-cell responses. It also produces complement proteins and cytokines essential for controlling *T. gondii* replication. Interferon-gamma (IFN-γ), in particular, is crucial for host defense, and reduced hepatic cytokine production can impair parasite containment, increasing systemic spread and neuroinvasion.

Based on observed associations in LHS, we hypothesize that liver dysfunction due to frailty, hepatitis, or alcoholism may further compromise these protective mechanisms, enabling *T. gondii* dissemination.

Immune dysregulation in AD patients was also evident in the LHS cohort, with significant reductions in key immune regulators, including vitamin D, T3, and folic acid. The bimodal distribution of folic acid suggests that both deficiency, which can impair immune function, and excessive supplementation, which may promote *T. gondii* replication via its reliance on folic acid metabolism, may influence parasite invasion.

## Conclusion

This large-scale, retrospective study, conducted over 20 years within a national health provider, is the first to identify a strong protective association between Ato/Pro and AD. Validation in the TriNetX U.S. network, encompassing over 120 million patients, reinforces the generalizability of these findings.

The persistence of risk reduction for over a decade post-treatment in both Israeli and American cohorts, alongside increased *T. gondii* seropositivity in AD patients, strongly suggests a link between toxoplasmosis elimination and AD protection. While causality cannot be definitively established, the consistent temporal associations across two large cohorts provide compelling support for the protective effect of Ato/Pro, with *T. gondii* involvement in AD pathogenesis being the most plausible explanation. Moreover, with Ato/Pro’s protective effect stronger in VZV-unvaccinated patients, we propose a unifying hypothesis wherein latent *T. gondii* infection, reactivated by herpesvirus infection in the context of immune dysfunction, impaired liver function, and neuroinflammation, ultimately drives AD. Future studies should further explore this mechanism and evaluate targeted interventions. Our findings suggest that controlled *T. gondii* elimination through antiprotozoals, potentially combined with agents that mitigate inflammation and prevent apoptosis in infected cells, such as selective or preferential COX-2 inhibitors, may represent a novel therapeutic avenue. Randomized controlled trials are needed to rigorously test this hypothesis and assess its clinical implications.

## Data Availability

Access to patients data is limited to researchers approved by the Institutional Review Board.

## Abbreviations

AD: Alzheimer’s Disease
Ato/Pro: Atovaquone/Proguanil
aOR: adjusted Odds Ratio
BBB: Blood-brain barrier
BMI: Body Mass Index
BP: Blood pressure
CNS: Central nervous system
COX-2: Cyclooxygenase-2
EHR: Electronic health records
FDR: False Discovery Rate
HCO: Healthcare organizations
HR: Hazards Ratio
ICD: International Classification of Diseases
IM: Intramuscular
LHS: Leumit Health Services
OR: Odds Ratio
PO: Per os
PSM: Propensity score matching
SES: Socioeconomic status
SMD: Standardized Mean Difference
T3: Triiodothyronine

## Footnotes

### Author contributions

Conceptualization: AI, EMe, Ema

Methodology: AI, EMe

Investigation: AI, SI, JS, Eme

Writing - original draft: AI

Writing - review & editing: AI, AW, SI, JS, SA, SV, EMa, EMe

## References

Contador I, Buch-Vicente B, del Ser T, Llamas-Velasco S, Villarejo-Galende A, Benito-León J, Bermejo-Pareja F. 2024. Charting Alzheimer’s Disease and Dementia: Epidemiological Insights, Risk Factors and Prevention Pathways. J Clin Med 13:4100. doi:10.3390/jcm13144100

Dominy SS, Lynch C, Ermini F, Benedyk M, Marczyk A, Konradi A, Nguyen M, Haditsch U, Raha D, Griffin C, Holsinger LJ, Arastu-Kapur S, Kaba S, Lee A, Ryder MI, Potempa B, Mydel P, Hellvard A, Adamowicz K, Hasturk H, Walker GD, Reynolds EC, Faull RLM, Curtis MA, Dragunow M, Potempa J. 2019. *Porphyromonas gingivalis* in Alzheimer’s disease brains: Evidence for disease causation and treatment with small-molecule inhibitors. Sci Adv 5. doi:10.1126/sciadv.aau3333

Flegr J. 2007. Effects of Toxoplasma on Human Behavior. Schizophr Bull 33:757–760. doi:10.1093/schbul/sbl074

Henriquez SA, Brett R, Alexander J, Pratt J, Roberts CW. 2009. Neuropsychiatric Disease and <i=Toxoplasma gondii</i= Infection. Neuroimmunomodulation 16:122–133. doi:10.1159/000180267

Hensley K. 2010. Neuroinflammation in Alzheimer’s Disease: Mechanisms, Pathologic Consequences, and Potential for Therapeutic Manipulation. Journal of Alzheimer’s Disease 21:1–14. doi:10.3233/JAD-2010-1414

Hlaváčová J, Flegr J, Fiurašková K, Kaňková Š. 2021. Relationship between Latent Toxoplasmosis and Depression in Clients of a Center for Assisted Reproduction. Pathogens 10:1052. doi:10.3390/pathogens10081052

Israel A, Merzon E, Schäffer AA, Shenhar Y, Green I, Golan-Cohen A, Ruppin E, Magen E, Vinker S. 2021. Elapsed time since BNT162b2 vaccine and risk of SARS-CoV-2 infection: test negative design study. The BMJ 375:e067873. doi:10.1136/bmj-2021-067873

Kovacs J. 1992. Efficacy of atovaquone in treatment of toxoplasmosis in patients with AIDS. The Lancet 340:637–638. doi:10.1016/0140-6736(92)92172-C

Kusbeci OY, Miman O, Yaman M, Aktepe OC, Yazar S. 2011. Could Toxoplasma gondii Have any Role in Alzheimer Disease? Alzheimer Dis Assoc Disord 25:1–3. doi:10.1097/WAD.0b013e3181f73bc2

Lathe R, Schultek NM, Balin BJ, Ehrlich GD, Auber LA, Perry G, Breitschwerdt EB, Corry DB, Doty RL, Rissman RA, Nara PL, Itzhaki R, Eimer WA, Tanzi RE. 2023. Establishment of a consensus protocol to explore the brain pathobiome in patients with mild cognitive impairment and Alzheimer’s disease. Alzheimer’s & Dementia 19:5209–5231. doi:10.1002/alz.13076

Ngoungou EB, Bhalla D, Nzoghe A, Dardé M-L, Preux P-M. 2015. Toxoplasmosis and Epilepsy — Systematic Review and Meta Analysis. PLoS Negl Trop Dis 9:e0003525. doi:10.1371/journal.pntd.0003525

Piotrowski SL, Tucker A, Jacobson S. 2023. The elusive role of herpesviruses in Alzheimer’s disease: current evidence and future directions. NeuroImmune Pharmacology and Therapeutics 2:253–266. doi:10.1515/nipt-2023-0011

Pomirchy M, Bommer C, Pradella F, Michalik F, Peters R, Geldsetzer P. 2025. Herpes Zoster Vaccination and Dementia Occurrence. JAMA. doi:10.1001/JAMA.2025.5013

Qiu C, Kivipelto M, von Strauss E. 2009. Epidemiology of Alzheimer’s disease: occurrence, determinants, and strategies toward intervention. Dialogues Clin Neurosci 11:111–128. doi:10.31887/DCNS.2009.11.2/cqiu

Readhead B, Haure-Mirande J-V, Funk CC, Richards MA, Shannon P, Haroutunian V, Sano M, Liang WS, Beckmann ND, Price ND, Reiman EM, Schadt EE, Ehrlich ME, Gandy S, Dudley JT. 2018. Multiscale Analysis of Independent Alzheimer’s Cohorts Finds Disruption of Molecular, Genetic, and Clinical Networks by Human Herpesvirus. Neuron 99:64–82.e7. doi:10.1016/j.neuron.2018.05.023

Silva MVF, Loures C de MG, Alves LCV, de Souza LC, Borges KBG, Carvalho M das G. 2019. Alzheimer’s disease: risk factors and potentially protective measures. J Biomed Sci 26:33. doi:10.1186/s12929-019-0524-y

Sutterland AL, Fond G, Kuin A, Koeter MWJ, Lutter R, van Gool T, Yolken R, Szoke A, Leboyer M, de Haan L. 2015. Beyond the association. Toxoplasma gondii in schizophrenia, bipolar disorder, and addiction: systematic review and meta analysis. Acta Psychiatr Scand 132:161–179. doi:10.1111/acps.12423

Taquet M, Dercon Q, Todd JA, Harrison PJ. 2024. The recombinant shingles vaccine is associated with lower risk of dementia. Nat Med 30:2777–2781. doi:10.1038/s41591-024-03201-5

Torrey EF, Bartko JJ, Yolken RH. 2012. Toxoplasma gondii and Other Risk Factors for Schizophrenia: An Update. Schizophr Bull 38:642–647. doi:10.1093/schbul/sbs043

Ukraintseva S, Yashkin AP, Akushevich I, Arbeev K, Duan H, Gorbunova G, Stallard E, Yashin A. 2024. Associations of infections and vaccines with Alzheimer’s disease point to a role of compromised immunity rather than specific pathogen in AD. Exp Gerontol 190:112411. doi:10.1016/j.exger.2024.112411

Wang Jianjun, Lin P, Li D, Yang B, Wang Jiaqi, Feng M, Cheng X. 2024. Analysis of the Correlation Between Toxoplasma gondii Seropositivity and Alzheimer’s Disease. Pathogens 13:1021. doi:10.3390/pathogens13111021

Wiener RC, Waters C, Bhandari R. 2020. The association of Toxoplasma gondii IgG and cognitive function scores: NHANES 2013-2014. Parasitol Int 78:102123. doi:10.1016/j.parint.2020.102123

Wyman CP, Gale SD, Hedges-Muncy A, Erickson LD, Wilson E, Hedges DW. 2017. Association between Toxoplasma gondii seropositivity and memory function in nondemented older adults. Neurobiol Aging 53:76–82. doi:10.1016/j.neurobiolaging.2017.01.018

Yang H-Y, Chien W-C, Chung C-H, Su R-Y, Lai C-Y, Yang C-C, Tzeng N-S. 2021. Risk of dementia in patients with toxoplasmosis: a nationwide, population-based cohort study in Taiwan. Parasit Vectors 14:435. doi:10.1186/s13071-021-04928-7

